# Extracellular vesicles proteomics-based machine-learning model predicts immunotherapy response in NSCLC

**DOI:** 10.64898/2026.03.12.26348220

**Authors:** Alejandro Castillo-Peña, Laura Boyero, Johana Cristina Benedetti, Amparo Sánchez-Gastaldo, Miriam Alonso, Miguel Ángel Muñoz-Fuentes, María Luisa Valdivia, Reyes Bernabé-Caro, Sonia Molina-Pinelo

## Abstract

Lung cancer remains the most common cause of cancer mortality worldwide. While the introduction of immune checkpoint inhibitors has changed the treatment landscape in non-small cell lung cancer, primary and acquired resistance continue to limits their long-term efficacy. The identification of reliable, minimally invasive biomarkers to guide immunotherapy response remains an urgent clinical need. In this study, we performed an in-depth proteomic profiling of plasma-derived small extracellular vesicles collected prior to treatment in 65 non-small cell lung cancer patients receiving pembrolizumab, aiming to uncover predictive molecular signatures. Mass spectrometry analysis initially detected over two thousand plasma extracellular vesicles proteins, including reported extracellular vesicle proteins, thereby validating our isolation and detection methodology. Among these, a four-proteins panel (MUC1, MUC5B, MUC5AC, and ANPEP) was associated with T cell dysfunction and an immunosuppressive tumor phenotype. This signature correlated with systemic inflammatory markers, particularly the platelet-to-lymphocyte ratio, and accurately predicted immunotherapy outcomes. Based on these findings, we stablished a plasma-derived score that strongly correlated with both progression-free and overall survival, providing a promising non-invasive tool for patient stratification and personalized immunotherapy.

## Introduction

Lung cancer is the leading cause of cancer-related mortality and the most frequently diagnosed cancer worldwide, accounting for one in eight cancer diagnoses and one in four cancer deaths ^1^. Most patients are diagnosed at advanced stage, contributing to poor survival outcomes ^2^. Non-small cell lung cancer (NSCLC) represents approximately 85% of lung cancer cases, with adenocarcinoma (ADC) and squamous cell carcinoma (SCC) as the predominant subtypes ^3–5^. Immune checkpoint inhibitors (ICIs) targeting the PD-1/PD-L1 axis, such as the anti-PD-1 antibody pembrolizumab, have revolutionized treatment of NSCLC and other tumor types ^6,7^. Despite their clinical success, only a subset of patients achieves durable responses, as primary or acquired resistance limits efficacy ^8,9^. This highlights the urgent need for reliable biomarkers to predict response and guide patient selection ^10^.

Current biomarkers, including PD-L1 expression, tumor mutational burden (TMB), and microsatellite instability (MSI), as well as systemic inflammatory markers such as the neutrophil-to-lymphocyte ratio (NLR), are limited by tumor heterogeneity, sampling invasiveness, cost, and lack of standardization ^11–14^. Liquid biopsy has emerged as a minimally invasive alternative that captures tumor heterogeneity and allows real-time monitoring of treatment response ^15,16^. Among liquid biopsy analytes, extracellular vesicles (EVs) are released by all cell types. They carry proteins, RNAs, and other molecules that reflect their cell of origin and modulate tumor-immune interactions ^17–24^. In tumor context, malignant cells release larger amounts of EVs, and their protein content mirrors the tumor proteome, offering the potential to uncover molecular signatures predictive of therapeutic response ^25^.

While most studies of EVs biomarkers have focused on RNA cargo, systematic analyses of the EVs proteome in NSCLC patients treated with immunotherapy remain scarce ^26–28^. ^25^Here, we set out to comprehensively characterize the baseline EVs proteome in NSCLC patients receiving pembrolizumab, with the aim of identifying protein-based biomarkers and developing a plasma-derived scoring system to enable non-invasive patient stratification and advance personalized immunotherapy.

## Materials and Methods

### Patient selection

The study included patients with advanced-stage NSCLC who were treated with pembrolizumab, either as monotherapy or in combination with chemotherapy, at the Virgen del Rocío University Hospital (Seville, Spain) between 2017 and 2023. Inclusion criteria required first-line immunotherapy with pembrolizumab, measurable tumors according to the Response Evaluation Criteria in Solid Tumors (RECIST) v1.1, absence of multiple primary tumors, and availability of a baseline plasma sample for the study. Of 274 patients available in the hospital database, 65 met all inclusion criteria (Fig. 1).

**Figure 1.**
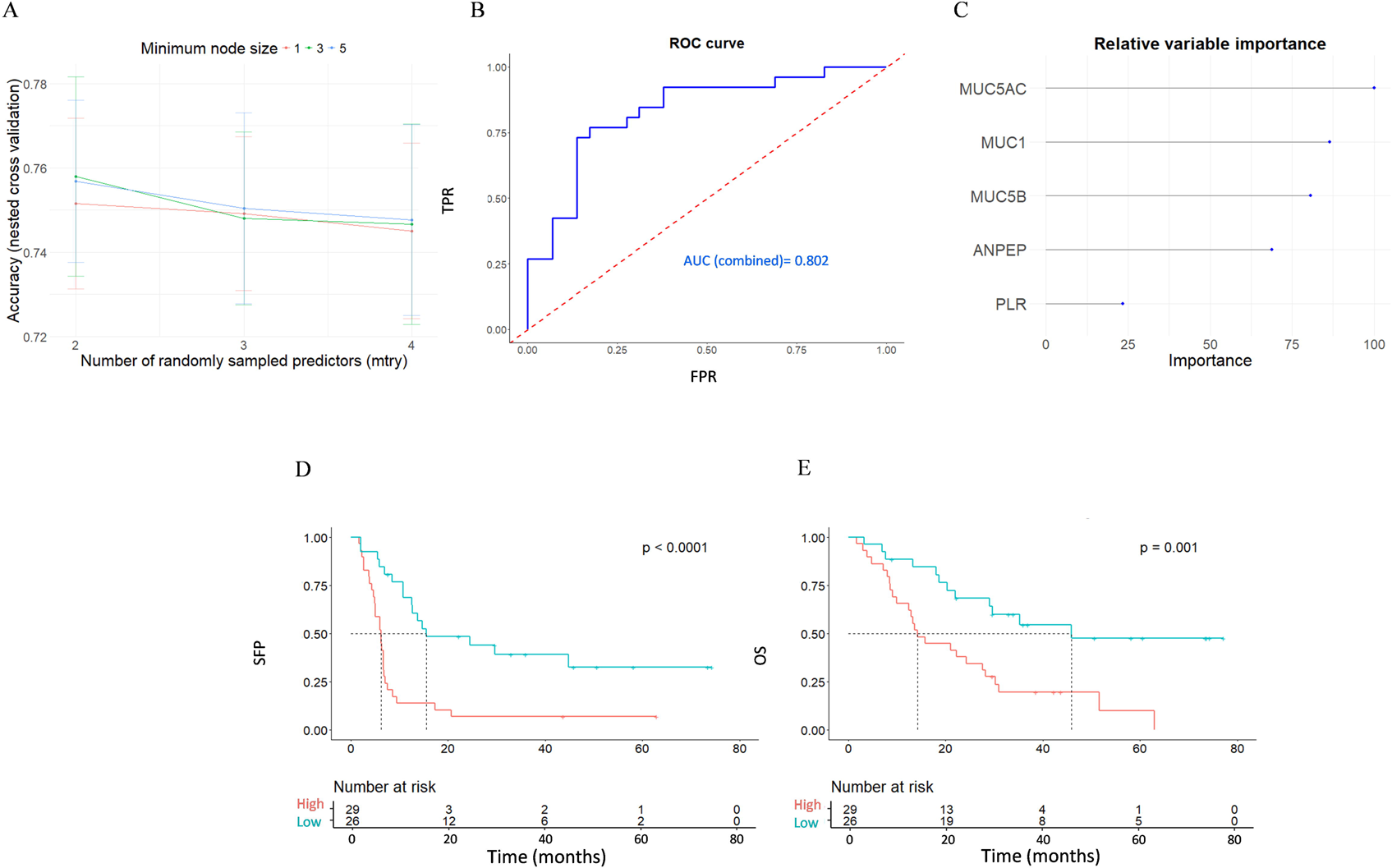
Flowchart of patient selection and final study cohort. Flow diagram ilustrating the screening and inclusion of NSCLC patients treated with immunotherapy at Virgen del Rocio University Hospital (HUVR) between 2017 and 2023. IT, immunotherapy; CR, complete response; PR, partial response; PD, progressive disease; SD, stable disease; NE, non-evaluable.

Treatment response was evaluated at six months according to RECIST v1.1. Patients with disease progression were classified as non-responders (NR), whereas those achieving partial or complete response were included in the responder group (R). The study was approved by the institutional ethics committee (SICEIA-2024-000105), and all participants provided informed consent through the institutional biobank.

Recorded clinical data included age, sex, histological subtype, smoking status, Eastern Cooperative Oncology Group (ECOG) performance status, disease stage, progression-free survival (PFS), overall survival (OS), and peripheral blood ratios. Neutrophil-to-lymphocyte ratio (NLR), monocyte-to-lymphocyte ratio (MLR), neutrophil plus monocyte-to-lymphocyte ratio (NMLR), and platelet-to-lymphocyte ratio (PLR) were calculated from routine complete blood counts.

### Sample collection and EVs isolation

Plasma samples were collected in 10 mL EDTA-coated tubes (BD Vacutainer). Tubes were centrifuged according to the manufacturer’s instructions to separate plasma, and samples were subsequently stored at −80 °C. Plasma-derived EVs were isolated using size exclusion chromatography (SEC) with 70 nm qEVoriginal Gen 2 columns (500 µl capacity; IZON), a method chosen for its ability to preserve vesicle integrity while minimizing co-isolation of plasma proteins. Plasma samples were first subjected to two consecutive centrifugation steps at 4 °C: 1,500 × g for 10 minutes to remove cellular debris, followed by 10,000 × g for 20 minutes to eliminate larger extracellular vesicles. Subsequently, 500 µl of pre-cleared plasma were loaded onto SEC columns, and five fractions of 400 µl each were collected following the manufacturer’s instructions. Fractions 1 and 5 were discarded, while fractions 2-4 were pooled and concentrated to a final volume of 50 µl in PBS using a SpeedVac system. All steps were performed gently to avoid vesicle disruption, and samples were kept on ice whenever possible.

### Nanoparticle Tracking Analysis (NTA)

The size distribution and concentration of extracellular vesicles were determined by NTA using a NanoSight Pro system (Malvern Panalytical, UK). EVs samples were previously isolated and diluted in freshly opened Gibco dPBS, filtered through a 0.22 µm membrane, and handled gently to minimize contamination with non-vesical particles. The analysis chamber was flushed with the same dPBS prior to each run. Measurements were acquired under a fixed focus of 2810, with an exposure time of 7.0 or 8.0 ms and contrast level set to 1. For each sample, five independent 60-second videos were recorded, and particle size distribution as well as and concentration curves were generated using the NAnoSight Pro analysis software (version 1.2.0.3).

### Transmission Electron Microscopy (TEM)

The morphology of isolated EVs was examined by TEM using a Zeiss Libra 120 transmission electron microscope (Carl Zeiss, Germany). Carbon-coated copper grids were pretreated by glow discharge to neutralize residual surface charges and increase hydrophilicity, promoting uniform adhesion of vesicles. Subsequently, 5 μL of the Evs suspension were placed on the grid for 20 min, followed by a 2-min wash with distilled water. Grids were negatively stained with 2% uranyl acetate for 10 min, excess stain was removed, and samples were air-dried in a desiccator before imaging.

### Protein extraction

Proteins from EVs samples, previously isolated and concentrated, were extracted using RIPA lysis buffer with protease inhibitors. Samples were then sonicated in a water bath for four cycles of 10 seconds each, keeping them on ice between cycles to prevent protein degradation.

### Inmunoblot

EVs identity was confirmed by Western blot analysis of canonical EVs markers. A total of 1 µg of protein was loaded per each gel lane. Specific antibodies against the tetranspanins CD9 (Immunostep, ref. 9PU-01MG) and CD63 (Thermo Fisher Scientific, ref. 10628D) were used as EVs markers. Reducing agents were omitted to preserve the native structure of these membrane proteins and avoid signal loss. To assess potential cellular contamination, calregulin (Santa Cruz biotechnology, ref. sc-373863) was used as a negative marker, using this time dithiothreitol (DTT) as a reducing agent. The secondary antibody used was an anti-mouse IgG produced in rabbit (Sigma Aldrich, ref. A9044).

### EVs proteomic analysis by Mass spectrometry

Previously to Mass spectrometry, protein precipitation was performed by sequentially adding 4 volumes of methanol, 1 volume of chloroform, and 3 volumes of Milli-Q water. After centrifugation (15,000 × g, 10 min, 4 °C) and removal of the aqueous phase, an additional 4 volumes of methanol were added. A second centrifugation step (15,000 × g, 10 min) was performed, the supernatant was discarded, and the samples were air-dried for 20 minutes at room temperature to remove any remaining liquid.

EVs protein samples were analyzed using a TimsTOF Bruker mass spectrometer (Bruker, Germany) operated in data-dependent acquisition (DDA-PASEF) mode. Initially, samples were dissolved in 8 M urea and 100 mM TEAB. After homogenization, proteins were reduced and alkylated with a final concentration of 5 mM tris(2-carboxyethyl)phosphine and 10 mM chloroacetamide at 37 °C under agitation for 1 h. Enzymatic digestion was then performed with trypsin (Promega) at 37 °C under agitation overnight, with an enzyme-to-protein ratio of 1:20. Resulting peptides were dried in a vacuum concentrator and desalted using Bond Elut C18 extraction cartridges (Agilent Technologies).

Peptides were subsequently injected onto a C18 analytical column (Aurora, 25 cm × 75 µm ID, IonOpticks, Australia) using a nanoElute 2 liquid chromatography system (Bruker Daltonics, Bremen, Germany) coupled to a TimsTOF Bruker SCP mass spectrometer. Peptides were eluded using a binary solvent system consisting of 0.1% formic acid in water (A) and 0.1% formic acid in acetonitrile (B), applying a linear gradient from 2% to 35% of solvent B at a flow rate of 250 nL/min. Data were acquired with controlled instrument parameters optimized for DDA-PASEF. All steps were performed at room temperature unless otherwise indicated, and care was taken to minimize sample loss and degradation.

### Differential Expression Analysis of EVs Proteins

Raw mass spectrometry data were processed using MaxQuant software (version 2.6.7.0) for peptide and protein identification and quantification. Database searching was performed with the Andromeda search engine integrated into MaxQuant. Relative protein quantification was performed using a label-free quantification (LFQ) approach, with the *Match between runs* feature enabled and a protein-level FDR threshold of 0.01. The UniProt Swiss-Prot human protein database, containing manually curated entries, was used to ensure high accuracy and consistency in protein identification.

Processed MaxQuant data were analyzed in R (version 4.3.2) using the DEP (Differential Enrichment analysis of Proteomics data) package, specifically designed for quantitative mass spectrometry data. This package enables filtering, normalization, imputation, and differential expression analysis using linear models implemented in the limma package.

Proteins annotated by MaxQuant as contaminants, reverse sequences, or identified only by site, as well as highly abundant plasma proteins (e.g., albumin and immunoglobulins), were removed prior to analysis. Peptide quality and the percentage of missing values per sample were assessed, and data normalization and imputation were performed. Differential expression analysis was then conducted and results were visualized using the R packages ggplot2, patchwork, and ggExtra.

### Functional Annotation and Pathway Enrichment of Identified EVs Proteins

The list of identified EVs proteins was compared with the 100 most commonly detected proteins in EVs obtained from Vesiclepedia ^29^. Functional annotation and pathway enrichment analyses were subsequently performed using the Database for Annotation, Visualization and Integrated Discovery (DAVID) Bioinformatics tool ^30,31^, integrating protein lists with biological functions and cellular processes. Kyoto Encyclopedia of Genes and Genomes (KEGG) pathway was used to identify relevant metabolic and signaling pathways, while Gene Ontology (GO) terms categorized protein by cellular components, molecular functions, and biological processes.

Graphical representations were generated using SRplot ^32^, an online platform that converts R scripts into complex visualtions, and the Reactome database ^33^ was used to create interactive maps of enriched pathways.

### Targeted Validation of Differentially Expressed Proteins by parallel reaction monitoring (PRM)

Validation of differentially expressed proteins was performed in a subset of 20 patients from the study cohort using a targeted proteomics assay based on Parallel Reaction Monitoring (PRM). A spectral library was first generated from DDA data of the study samples to select candidate peptides for the proteins of interest, using MSFragger v4.3 within the FragPipe v23.1 pipeline. The resulting .pepXML files were imported into Skyline v25.1, which was employed for chromatogram visualization and quantitative analysis of PRM data. Oxidation (M) and N-terminal acetylation were considered as variable modifications, while carbamidomethylation (C) was set as a fixed modification. PRM analyses were performed on a timsTOF SCP mass spectrometer operated under the same chromatographic conditions used for DDA acquisition. Only proteotypic peptides meeting the following quality criteria were quantified: dotp and idotp ≥ 0.75, at least four detected fragment transitions, and a retention time deviation within ±2 min. Peak area intensities were normalized using total ion current (TIC), log₂-transformed, and subsequently analyzed statistically in R. Quantification was performed by summing the MS1 and MS2 signal intensities.

### Predictive modeling of Immunotherapy Response

To evaluate the predictive capacity of differentially expressed proteins between R and NR, a Random Forest classification model was developed in R (version 4.3.2) using the caret package (Classification And Regression Training) for model training and validation, and the ranger package for optimized Random Forest implementation. A random seed was set to ensure reproducibility.

Input data included the response variable (R vs NR) and intensity values of the differentially expressed EVs proteins. The Random Forest model was trained using 1000 trees, while the remaining hyperparameters were optimized through a tuning procedure.

Model validation was performed using two complementary approaches: bootstrapping and repeated K-fold cross-validation. Variable importance was assessed using the Gini impurity metric, and rankings from bootstrapping and cross-validation were compared to identify the most predictive proteins. The top-ranked proteins were then used to construct a simplified final model, built using nested cross-validation, in which the inner loop was employed for hyperparameter tuning and the outer loop for unbiased performance evaluation.

Model performance was evaluated using accuracy, Kappa coefficient, sensitivity, specificity, and area under the ROC curve (AUC). ROC curves were plotted using the ROCR package in R.

### Statistical and Bioinformatic Analysis

Clinical variables were analyzed separately for R and NR, and hypothesis testing were conducted to identify potential significant differences between groups. Categorical variables were evaluated using the chi-square test or Fisher’s exact test, while numerical variables were assessed using the t-test or Wilcoxon rank-sum test, depending on whether the data are normally distributed. Qualitative evaluation of PRM protein detection was also carried out using Fisher’s exact test. Correlation of Differentially Expressed EVs Proteins with peripheral blood ratios were evaluated in R (version 4.3.2), with normality assessed using Shapiro-Wilk test. Pearson’s correlation was applied to normally distributed data, and Spearman’s rank correlation for non-parametric data. Patient survival was analyzed using the survival and survminer packages. Progression-free survival (PFS) was defined as the time from treatment initiation to either disease progression or death, while overall survival (OS) was defined as the time from treatment initiation to death from any cause. Patients were stratified into high- and low-expression groups based on the Youden index, Kaplan–Meier survival estimates were generated, and with group comparisons performed using the log-rank test. The contribution of smoking status as a covariate to protein expression differences was assessed using linear regression models implemented in R (stats::lm).

The risk index was calculated from identified EVs protein intensity values using a penalized regression model based on the Elastic Net method, implemented with the glmnet package. This model combines Lasso and Ridge penalties to balance variable selection with coefficient shrinkage in the presence of collinearity, improving interpretability and robustness, reducing overfitting, and enhancing generalizability. Continuous risk scores generated by the model were classified into high- and low-risk groups using the Youden index. Finally, correlation data between protein expression in the model and T cell dysfunction were obtained from TIGER (Tumor Immunotherapy Gene Expression Resource) ^34,35^. In this resource, T cell dysfunction was inferred from predefined gene signatures associated with this immunosuppressive mechanism ^35^.

## Results

### Patient Characteristics and Clinical Outcomes

The study cohort included 65 patients (32 R, 33 NR) whose clinical characteristics are summarized in Table 1. Demographics and clinical features were broadly comparable between groups, with smoking status as the only clinical variable showing a significant imbalance. NR also displayed higher baseline systemic inflammatory ratios (NLR, NMLR, and PLR) and significantly shorter PFS and OS than responders.

**Table 1.** Clinical characteristics of patients included in the baseline proteomic analysis.

| Characteristics | Full cohort n=65 | Responders n=32 (49.2%) | Non-responders n=33 (50.8%) | p-value |
| --- | --- | --- | --- | --- |
| Age (years) | 67 [60-75] | 70 (64-76) | 65 (58-69) | 0.080 |
| Sex, n (%) |  |  |  | 0.303 |
| Male | 55 (84.6) | 29 (90.6) | 26 (78.8) |  |
| Female | 10 (15.4) | 3 (9.4) | 7 (21.2) |  |
| Histological subtype, n (%) |  |  |  | 0.674 |
| Adenocarcinoma | 47 (72.3) | 23 (71.9) | 24 (72.7) |  |
| Squamous cell carcinoma | 10 (15.4) | 6 (18.8) | 4 (12.1) |  |
| NA | 8 (12.3) | 3 (9.4) | 5 (15.2) |  |
| Smoking status, n (%) |  |  |  | 0.004 |
| Never smoker | 6 (9.2) | 0 (0.0) | 6 (18.2) |  |
| Former smoker (+10 years) | 15 (23.1) | 11 (34.4) | 4 (12.1) |  |
| Former smoker (-10 years) | 17 (26.2) | 11 (34.4) | 6 (18.2) |  |
| Current smoker | 27 (41.5) | 10 (31.3) | 17 (51.5) |  |
| ECOG, n (%) |  |  |  | 0.249 |
| 0-1 | 61 (93.8) | 31 (96.9) | 30 (90.9) |  |
| 2 | 4 (6.2) | 1 (3.1) | 3 (9.1) |  |
| Stage |  |  |  | 0.200 |
| I | 2 (3.1) | 2 (6.3) | 0 (0.0) |  |
| II | 1 (1.5) | 0 (0.0) | 1 (3.0) |  |
| III | 9 (13.8) | 6 (18.8) | 3 (9.1) |  |
| IV | 53 (81.5) | 24 (75.0) | 29 (87.9) |  |
| Treatment |  |  |  | 0.375 |
| Pembrolizumab | 36 (55.4) | 20 (62.5) | 16 (48.5) |  |
| Combined immunotherapy | 29 (44.6) | 12 (37.5) | 17 (51.5) |  |
| MLR | 0.44 (0.35-0.58) | 0.41 (0.35-0.52) | 0.50 (0.36-0.69) | 0.189 |
| NLR | 3.6 (2.7-5.0) | 3.0 (2.4-4.7) | 4.2 (3.2-6.1) | 0.038 |
| NMLR | 4.1 (3.1-5.6) | 3.4 (2.8-5.1) | 4.8 (3.6-6.7) | 0.038 |
| PLR | 163.1 (125.6- | 135.4 (91.3-184.3) | 194.4 (148.0-285.8) | 0.007 |
| Progression-Free Survival (months) | 7.1 [5.0-16.4] | 20.4 [12.2-44.0] | 5.0 (3.1-6.4) | 3.28E-11 |
| Overall survival (months) | 22.3 [10.0-36.9] | 33.5 [20.3-53.3] | 13.3 (8.1-27.4) | 1.24E-04 |

### Comprehensive Characterization and Proteomic Profile of EVs

Plasma-derived EVs from NSCLC patients treated with pembrolizumab were successfully isolated and characterized using complementary approaches. Western blot confirmed enrichment of the EV markers CD9 and CD63 and absence of the cellular contamination calregulin (Fig. 2A). TEM validated the presence of EVs within the characteristic size range of small EVs (30–150 nm) with typical cup-shaped morphology (Fig. 2B). NTA further confirmed a predominant size peak around 100 nm and particle concentrations of ∼10¹¹ particles/mL (Fig. 2C), consistent with an small EVs-enriched preparation.

**Figure 2.**
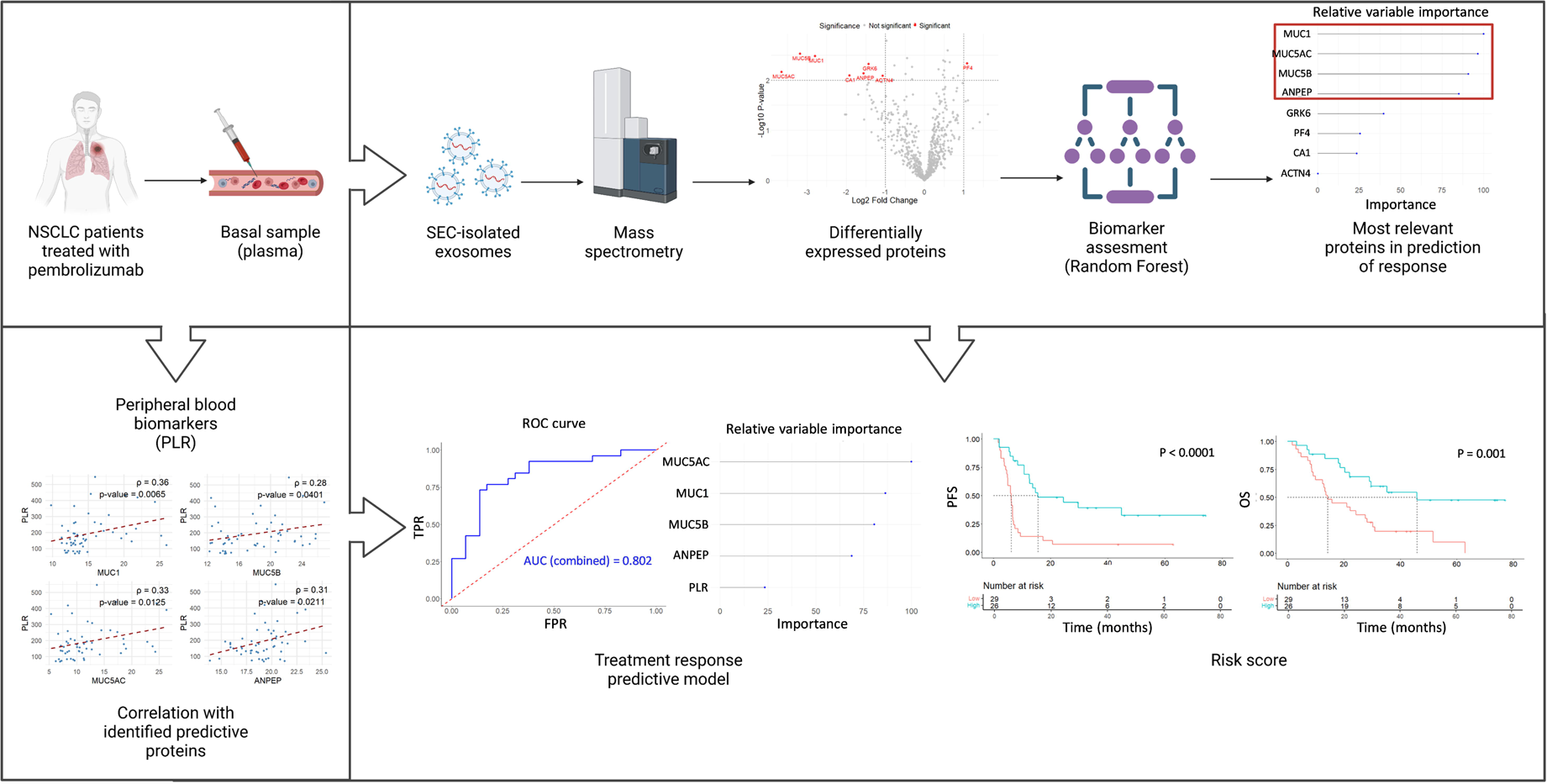
Characterization of plasma-derived EVs from NSCLC patients treated with pembrolizumab and their protein content. **A)** Western blot analysis showing enrichment of EV markers CD9 and CD63 and absence of the cellular marker calregulin, **B)** Representative transmission electron microscopy image of small EVs displaying the expected morphology and size range, **C)** Nanoparticle tracking analysis representative of all analyzed samples, showing size distribution and particle concentration, **D)** Overlap of protein identification across samples, represented as the number of proteins shared among increasing number of samples, **E)** Venn diagram comparing the 100 most representative EV proteins reported in Vesiclepedia with the total set of proteins identified (left) and with the remaining proteins after the filtering process, **F)** KEGG pathway enrichment analysis. The X-axis shows enrichment scores (–log₁₀ FDR), dot size represents the number of proteins per pathway, and color indicates statistical significance, and **G**) Gene Ontology enrichment analysis for Biological Process (BP), Cellular Component (CC), and Molecular Function (MF).

Following protein extraction and mass spectrometry, data were processed using MaxQuant. One sample was excluded due to incorrect processing, leaving 64 samples for analysis. Proteins classified as contaminants, reverse sequences, highly abundant plasma proteins (e.g., albumin and immunoglobulins), or identified solely by site-specific modifications without unique peptides were excluded. In addition, samples with more than 60% missing values, defined as the proportion of proteins with zero intensity relative to the total number of identified proteins across all samples, were excluded (Fig. S1A), resulting in a final dataset of 55 samples. Proteins were then filtered to retain only those proteins with valid values in at least 80% of samples from one or both study groups, reducing the dataset from 2,267 to 720 proteins. Among these, all patients had valid intensity values for more than 600 proteins, and approximately 400 proteins were identified in all samples (Fig. 2D). Peptide distributions by length (7–30 amino acids) and mass-to-charge ratio (m/z) (between 400–1200) were consistent with efficient fragmentation during mass spectrometry (Fig. S1B). The final dataset (n = 55) was normalized using the variance stabilization (VSN) method and missing values were inferred to follow MNAR pattern. Quantile regression imputation of left-censored data (QRILC) was applied, with no significant alterations in overall data distribution observed (Fig. S1C-E).

Functional enrichment analysis of the 720 filtered proteins confirmed their biological relevance to EVs, lung cancer, and immune response. To assess whether the proteomic profile of our samples reflected the canonical extracellular vesicle composition, identified proteins were compared with the 100 most frequently reported EV proteins in Vesiclepedia, a manually curated database compiling molecular content data in EVs ^29^. Of these, 99 were present in the full dataset of 2,267 proteins, and 94 persisted after filtering (Fig. 2E). These findings indicate that the majority of the identified proteins correspond to the typical EV protein repertoire, suggesting that the isolation procedure was properly conducted. KEGG pathway enrichment highlighted actin cytoskeleton regulation, leukocyte transendothelial migration, proteoglycans in cancer, and signaling pathways such as RAP1 (Fig. 2F), all of which are involved in processes associated with extracellular vesicles and the tumor microenvironment. GO terms related to cytoskeleton organization, extracellular exosome, and GTPase activity were significantly enriched (Fig. 2G). Finally, Reactome analysis generated an interactive map of enriched biological pathways, highlighting processes including immune response, vesicle-mediated transport, cell–cell communication, autophagy, and programmed cell death (Fig. S2).

### EVs proteomic differences between immunotherapy R and NR

EVs proteomes from R and NR to pembrolizumab were compared to identify proteins differentially expressed between the two groups. Using normalized and imputed protein intensity data, differential expression analysis identified eight proteins significantly differentially expressed between R and NR (p < 0.01, fold change (FC) >2 or < −2): MUC1, MUC5B, MUC5AC, ANPEP, ACTN4, CA1, GRK6, and PF4 (Fig. 3A). PCA based on these eight proteins showed a clear segregation of samples according to clinical response to pembrolizumab (Fig. 3B). Among the eight differentially expressed proteins, seven (MUC1, MUC5B, MUC5AC, ANPEP, CA1, ACTN4, GRK6) were downregulated in R relative to NR, whereas PF4 was upregulated (Fig. 3C). The three mucins MUC1 (FC = −6.964), MUC5B (FC = −9.063) and MUC5AC (FC = −12.553) showed the largest relative expression changes between groups (Fig. 3D).

**Figure 3.**
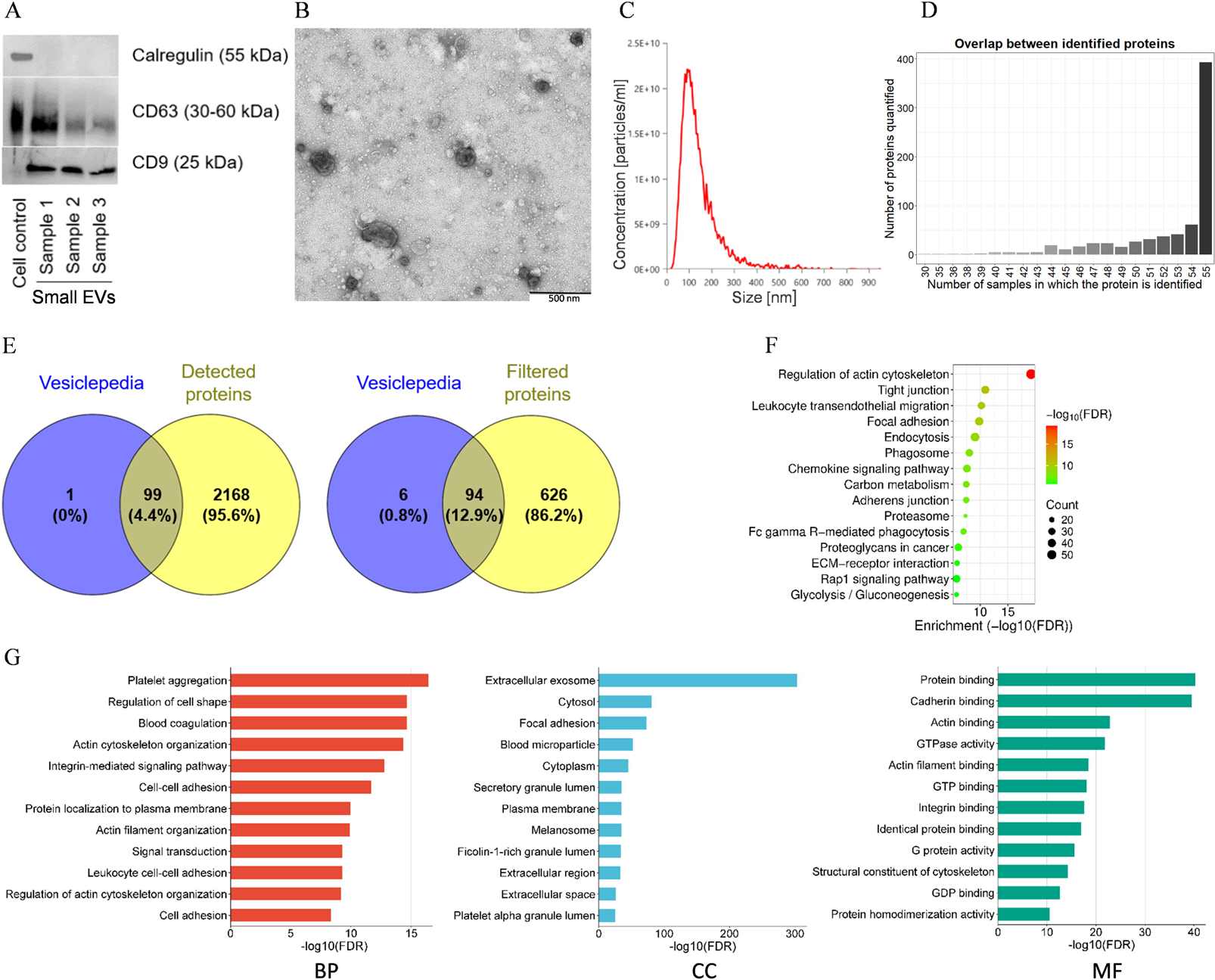
Differential expression analysis of EV proteins. **A)** Volcano plot showing differentially expressed proteins between R and NR, with significance threshold of < 0.01 and a fold change > 2 or < - 2, **B)** PCA of the 8 proteins exhibiting the largest variability between R and NR, **C)** Boxplot of protein intensities, with each dot representing an individual sample, color-coded by patient within each response group, and **D)** Bar plot depicting fold change (FC) for each protein. Blue bars indicate lower expression in R relative to NR, and red bars indicate higher expression. **E)** Adjusted expression difference and 95% CI, estimated using linear models including smoking status as a covariate. The p-value corresponds to the effect of treatment response for each protein.

Among the clinical variables considered in this study, the only one that differed significantly between R and NR was smoking status. To evaluate wether this variable could be acting as a confounder in the differential expression analysis, a separate linear model was fitted for each protein including smoking status as a covariate. To improve statistical power, patients were grouped into a binary variable based on tobacco exposure: recent tobacco exposure, which included current smokers and former smokers who quit < 10 years ago; and no recent tobacco exposure, which included former smokers who quit ≥ 10 years ago and never smokers. The eight proteins remained significantly differentially expressed between R and NR independently of smoking status (Fig. 3E), whereas no statistically significant associations attributable to smoking status were observed for most of them (MUC1, p = 0.460; MUC5B, p = 0.180; MUC5AC, p = 0.134; ANPEP, p = 0.358; ACTN4, p = 0.342; CA1, p = 0.064; GRK6, p = 0.273), except of PF4 (p = 0.030). In any case, for some proteins a slightly degree of separation in the distribution of intensities within R and NR according to tobacco exposure can be observed (Fig. S3).

### Prognostic impact of differentially expressed EVs proteins

Proteins differentially expressed between R and NR were further evaluated for their potential prognostic value. The optimal cut-off to define high and low expression groups for each protein was determined using the Youden index. Five proteins showed a significant association with PFS: MUC1 (HR = 0.264, 95% CI = 0.135-0.517, p < 0.0001), MUC5B (HR = 0.453, 95% CI = 0.245-0.837, p = 0.010), MUC5AC (HR = 0.513, 95% CI = 0.280-0.940, p = 0.029), ANPEP (HR = 0.481, 95% CI = 0.262-0.882, p = 0.016), and PF4 (HR = 2.115, 95% CI = 1.113-4.016, p = 0.020). The remaining three proteins did not show a significant association with PFS survival: ACTN4 (HR = 0.587, 95% CI = 0.317-1.088, p = 0.088), CA1 (HR = 0.562, 95% CI = 0.294-1.077, p = 0.080), and GRK6 (HR = 0.542, 95% CI = 0.280-1.050, p = 0.066) (Fig. 4).

**Figure 4.**
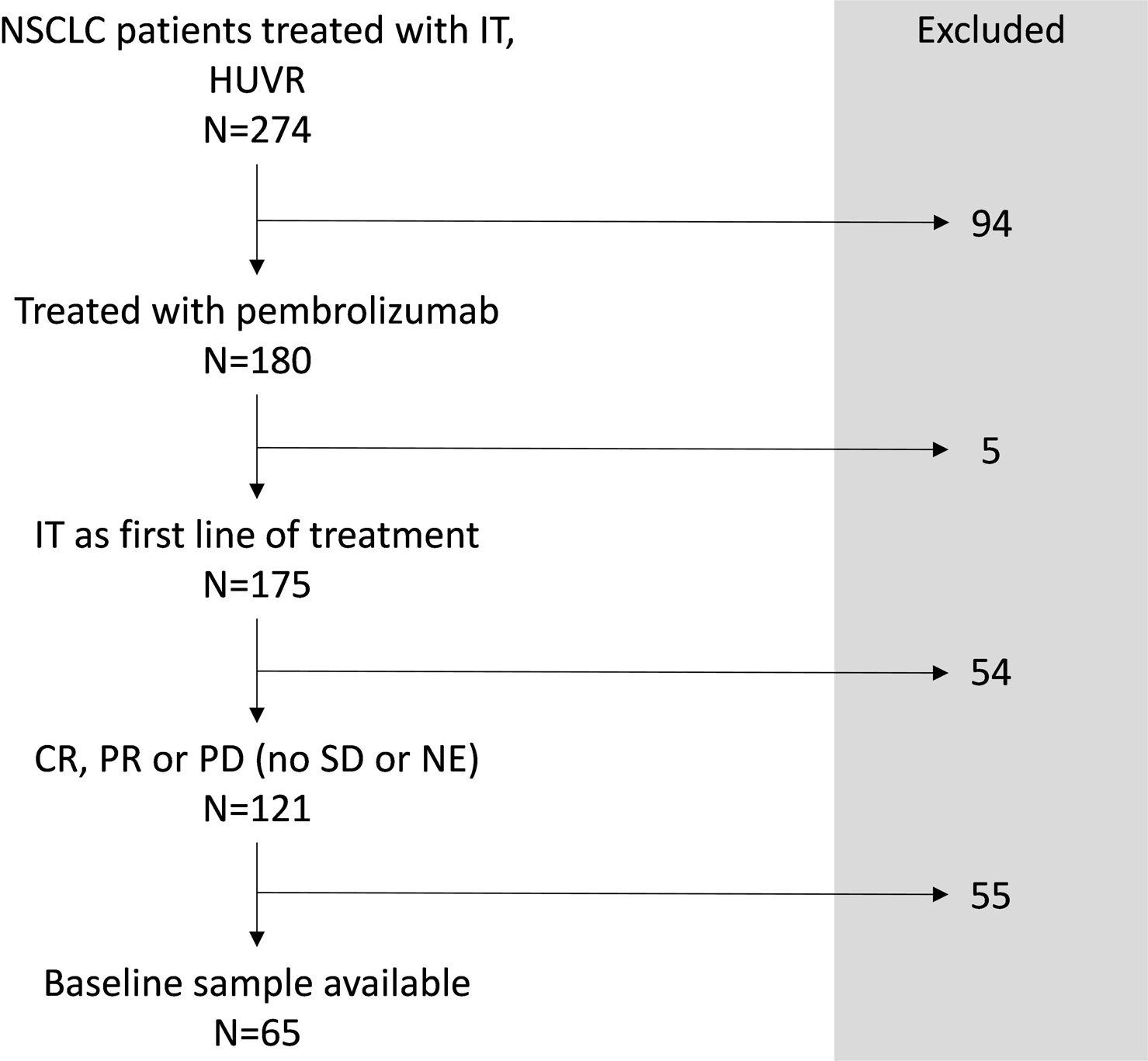
Progression-free survival (PFS) analysis of differentially expressed EV proteins. A-H) Kaplan–Meier curves show PFS stratified by high and low protein expression, with cut-off points determined using the Youden index. **A)** MUC1, **B)** MUC5B, **C)** MUC5AC, **D)** ANPEP, **E)** PF4, **F)** ACTN4, **G)** CA1, **H)** GRK6. P-values are indicated. **I)** Forest plot showing the hazard ratios (HR) and 95% confidence intervals derived from analyzed EV proteins. Red dots correspond to significant associations with HR < 1, blue dots to HR > 1, and black dots to proteins whose confidence interval includes HR = 1.

In contrast, only ACTN4 (HR = 0.511, 95% CI = 0.266-0.981, p = 0.04) showed a significantly association with OS, whereas none of the remaining seven proteins showed a significant correlation when analyzed individually (Fig S4).

### EVs proteome-based prediction of immunotherapy response

To identify which of the eight candidate proteins has the highest predictive potential for immunotherapy response, a Random Forest classification model was constructed. This approach estimates the relative importance of each variable based on its contribution to the overall predictive performance. Given the limited cohort size and the absence of an external validation cohort, repeated 5-fold cross-validation was used for internal validation. The dataset was partitioned into five folds of 11 samples each, and the process was repeated 10 times, generating 50 distinct training and validation sets. The model was built with 1000 trees, a minimal node size of 3, and 2 randomly selected predictors per tree (Fig. 5A). The model achieved a mean accuracy of 0.752 (95% CI = 0.545–0.979), a Kappa coefficient of 0.501, a sensitivity of 0.713, a specificity of 0.787, and an AUC of 0.757 (Fig. 5B), demonstrating predictive performance notably better than randomness in predicting the response to immunotherapy. The four proteins contributing most to the model were the three mucins (MUC1, MUC5B, MUC5AC) and ANPEP, all with relative importance values exceeding 85% (Fig. 5C). To verify stability and rule out overfitting, a second Random Forest model using bootstrapping as the resampling method produced highly concordant variable importance rankings, confirming that these four proteins consistently occupied the top positions (Fig. 5D).

**Figure 5.**
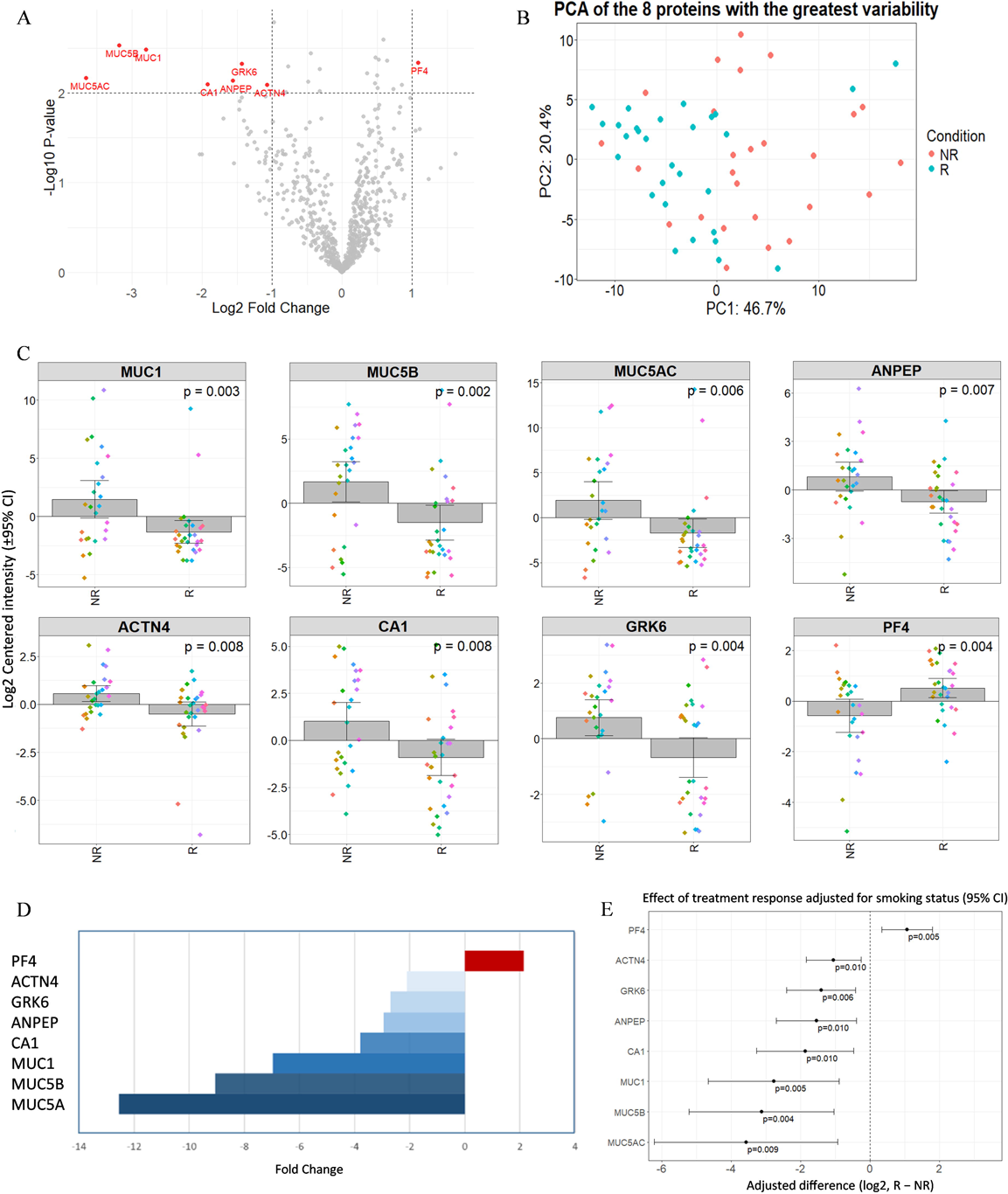
Random Forest model predicting immunotherapy response based on EVs proteomic profiles. **A)** Mean accuracy of the model across repeated 5-fold cross-validation for different hyperparameter combinations. The configuration with the highest performance was selected. **B)** ROC curve of the final model. **C)** Relative importance of each protein in the final model, calculated using the Gini impurity metric. **D)** Comparison of protein importance rankings between the models validated by bootstrapping (red) and repeated cross-validation (blue). **E)** Spearman correlation between the four top-ranked predictive proteins (MUC1, MUC5B, MUC5AC, and ANPEP) and the PLR ratio. **F)** Heatmap showing the correlation between the expression of the proteins included in the model and T cell dysfunction, inferred from the expression of gene signatures associated with T cell dysfunction. Data were obtained from TIGER.

To explore the potential biological relevance of the differentially expressed EVs proteins in NSCLC resistance to pembrolizumab, we examined their correlation with peripheral blood biomarkers that have previously reported as predictive or prognostic value in NSCLC patients treated with immunotherapy, such as the NLR, MLR, NMLR and PLR. All four proteins showed a significant positive correlation with PLR (Fig. 5E). No significant correlations were found between the proteins under study and the MLR or NLR ratios, despite positive coefficients (Fig. S5). Given the interest in exploring the relationship between these proteins and the immune system, we evaluated the correlation of the four proteins with T cell dysfunction in the two main NSCLC subtypes using gene expression data from TIGER, a bioinformatics platform that integrates large-scale transcriptomic datasets from immunotherapy-treated patients and allows exploration of predefined gene signatures across diverse tumor contexts. Interestingly, this correlation was positive and significant for all four proteins in SCC, but only for MUC1 and ANPEP in ADC. ANPEP showed a stronger correlation with T cell dysfunction than the mucins in both histological subtypes (Fig. 5F).

### PRM-based validation of candidate proteins

To validate the differential expression observed in the four proteins with the highest predictive potential (MUC1, MUC5B, MUC5AC, and ANPEP), a targeted PRM proteomics analysis was performed. Among the analyzed peptides, only those that met the predefined quality parameters in a sufficient number of samples were quantified; these are listed in Table S1. Peptide quantification was based on the sum of integrated MS1/MS2 peak areas, and protein abundance was calculated as the median intensity of all corresponding peptides. The expected trend was observed for all analyzed peptides (Fig. 6A), with a significant overexpression in non-responders (NR) for MUC1 (log_2_FC = 3.56, p = 1.35×10^-4^), MUC5B (log_2_FC = 4.01, p = 5.52×10^-5^) and ANPEP (log_2_FC = 2.59, p = 0.002). In the case of MUC5AC, its target peptides were consistently detected in NR but not in R (p = 0.011), suggesting that its expression levels in responders were close to the detection threshold, while showing marked overexpression in NR. Therefore, due to the absence of signal in R, no fold change could be estimated (Fig 6B).

**Figure 6.**
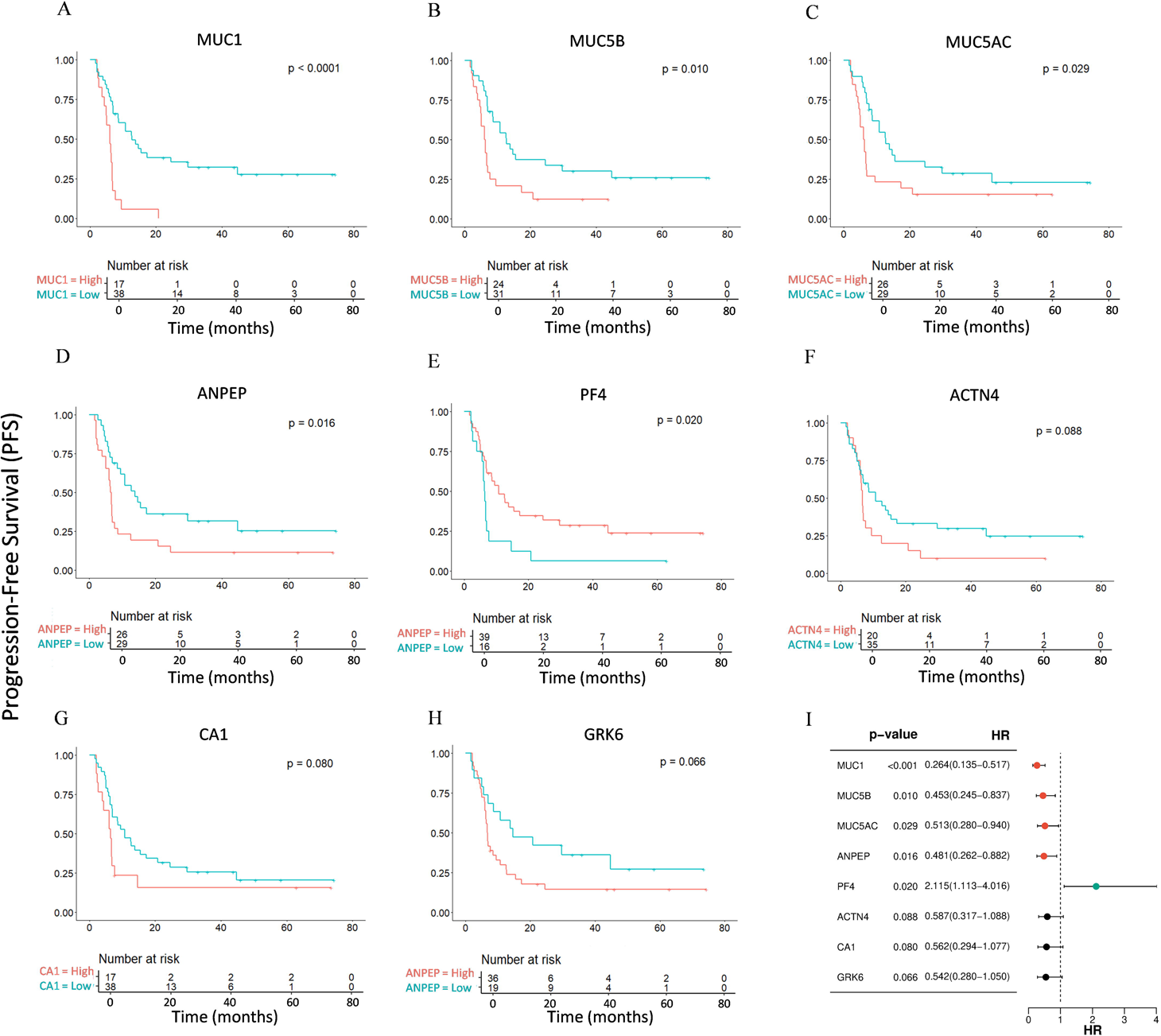
PRM analysis of candidate proteins. **A)** Boxplots showing the intensities of the quantified peptides, and **B)** boxplots of protein intensities, calculated as the median of their corresponding peptides, log₂-normalized, and grouped by clinical condition. Each point represents an individual sample.

### EVs protein- and PLR-based risk predicts immunotherapy response

To assess the clinical utility of the predictive biomarkers, a refined Random Forest model was built using the four proteins identified in the initial analysis (MUC1, MUC5B, MUC5AC, and ANPEP) together with the PLR ratio as a fifth predictor. Model robustness was evaluated using nested cross-validation, with a 10-fold cross-validation repeated 10 times in the inner loop for hyperparameter optimization (Fig. 7A), and a 10-fold cross-validation in the outer loop for unbiased performance estimation. The final model achieved a mean accuracy of 0.787 (95% CI, 0.687–0.886), a mean Kappa coefficient of 0.585, a sensitivity of 0.800, a specificity of 0.800, and a combined AUC of 0.802 (Fig. 7B), indicating improved discriminative ability and stability compared to the initial model. Although PLR contributed to predictive performance, its relative importance was lower than that of the EVs proteins (Fig. 7C). The remaining clinical variables were also evaluated using a Random Forest model, but they were excluded from the final model because their relative importance in prediction was notably lower than that of the proposed EVs proteins (Fig. S6). To translate these finding into a clinically applicable tool, the five predictors were combined into a continuous risk score using an Elastic Net model. Patients were classified into high- and low-risk groups for non-response to immunotherapy based on the Youden index. Kaplan-Meier analysis revealed that high-risk patients had significantly shorter PFS (HR = 0.284, 95% CI = 0.149–0.540, and p < 0.0001) and OS (HR = 0.333, 95% CI = 0.165–0.671, and p = 0.001) compared to low-risk patients (Fig. 7D-E). These results demonstrate that the combined EVs protein and PLR panel provides a robust predictive and prognostic tool, outperforming individual predictors and offering potential clinical utility for guiding pembrolizumab therapy in NSCLC patients.

**Figure 7.**
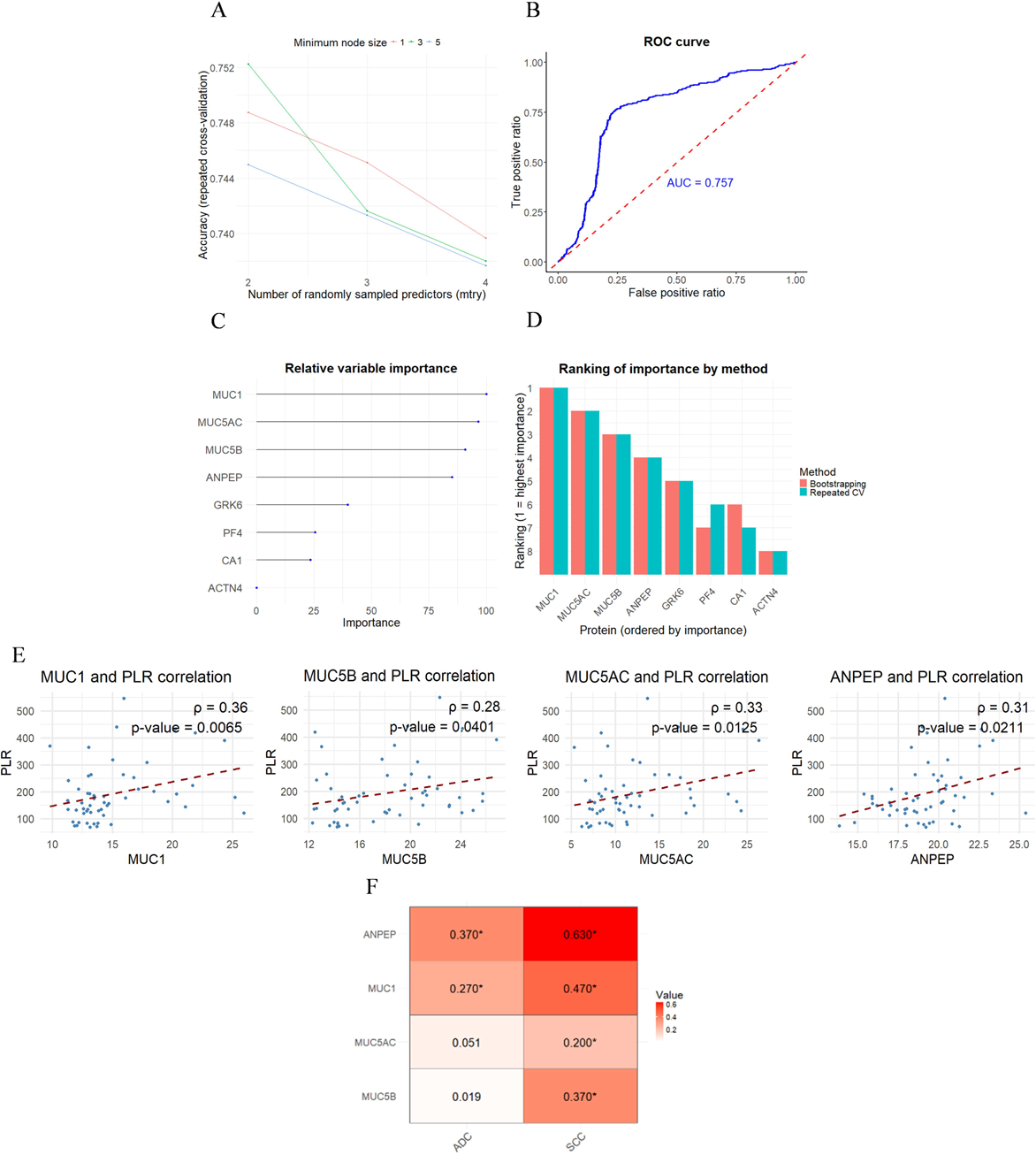
Optimized Random Forest model risk stratification of NSCLC patients based on the four identified proteins and PLR. **A)** Mean accuracy of different hyperparameter configurations evaluated by repeated cross-validation in the inner loop. Bars indicate the standard deviation across iterations. The best-performing set (mtry = 2, minimum node size = 3) was selected. **B)** ROC curve of the final model, showing the combined AUC from the 10 iterations of the outer loop. **C)** Relative importance of each predictor in the final model, calculated using the Gini impurity index. **D)** Progression-free survival and **E)** overall survival of patients stratified into high- and low-risk groups according to the Elastic Net-derived risk score.

## Discussion

This study provides the first in depth analysis of the plasma-derived EVs proteome in NSCLC patients treated with pembrolizumab, with the goal of identifying protein signatures that could predict treatment outcome. Using mass spectrometry, we detected more than 2,200 proteins, of which 720 met the criterion of valid measurements in at least 80% of samples and included reported markers of extracellular vesicle, which supports the robustness of our isolation and detection workflow. Pathway enrichment analyses further revealed processes related to tumor progression, underscoring that EVs protein cargo reflects the biological state of the tumor and its potential evolution.

Although the overall EVs proteome was broadly similar between R and NR, we identified eight proteins (MUC1, MUC5B, MUC5AC, ANPEP, ACTN4, CA1, GRK6, and PF4) that were differentially expressed. Seven of these were more abundant in NR, which could reflect higher levels of synthesis, increased vesicle release, or a selective packaging into EVs. When smoking status was included as a covariate, all eight proteins remained significantly associated with treatment response after adjustment, whereas smoking status was not significant except for PF4, suggesting a possible additional or concomitant effect. In some cases, a trend toward higher protein levels with recent tobacco exposure was observed, warranting further study on wether smoking contributes to therapy resistance. Among the eight proteins, MUC1, MUC5B, MUC5AC, ANPEP, and PF4 showed significantly association with PFS but not with OS, suggesting that they may primarily influence early responses to immunotherapy and contribute to primary resistance ^9,36^. In contrast, ACTN4 was the only protein associated with OS, consistent with its known role in cytoskeletal regulations and tumor progression ^37^. Notably, ACTN4 expression has been associated with brain metastasis and found to be higher in metastatic compared with primary lung cancer tissue ^38^. Its reported predictive value for Osimertinib response in EGFR-mutated tumors, an immune-cold phenotype with limited responsiveness to immunotherapy, supports its role as a general marker of poor prognosis in NSCLC ^39,40^. Taken together, these findings indicate that no single protein robustly predicts both PFS and OS, underscoring the need for composite biomarker signatures to achieve clinically meaningful prognostic value.

Among these proteins, MUC1, MUC5B, MUC5AC and ANPEP emerged as the strongest predictors of immunotherapy response, clearly outperforming all clinical variables analyzed. These proteins have been reported to be implicated in lung carcinogenesis and immune modulation ^41,42^. Specifically, MUC1, MUC5AC, and MUC5B are predominantly overexpressed in lung adenocarcinoma, contributing to tumor aggressiveness and adverse clinical outcomes ^42–44^. *In vitro*, MUC1 upregulates PD-L1 in tumor cells and PD-1 in CD8+ T cells ^45–47^, while inhibiting T cell proliferation and activation, ultimately leading to depletion of tumor-infiltrating lymphocytes ^48,49^. Through these mechanisms, MUC1 impairs immune recognition and elimination of tumor cells. Similarly, MUC5B promotes immune evasion via the Wnt/β-catenin signaling pathway, enhancing PD-1 and PD-L1 expression while suppressing CD8+ T cell activity, thereby fostering an immunosuppressive tumor microenvironment ^50^. Although the role of MUC5AC in cancer immunotherapy is less understood, it has been associated with immunosuppressive signaling and tumor aggressiveness through multiple pathways ^51–53^.

ANPEP, also known as CD13, is involved in innate immunity and inflammation, with overexpression linked to angiogenesis, tumor growth, and metastasis in several tumors, including lung cancer ^41,54,55^. It has been identified as a surface marker of malignant myeloid cells ^41^. Previous studies reported a positive correlation between ANPEP and PD-1 expression, and PD-1 inhibition reduce CD13^+^ myeloid cells in NSCLC patients ^56,57^. Building on these observations, we found that ANPEP and the mucins correlate with T cell dysfunction, supporting their contribution to an immunosuppressive tumor microenvironment, primarily through inactivation of antitumor immune cells. However, functional validation will be required to confirm these mechanisms.

We also analyzed the correlation of these four proteins and established peripheral blood biomarkers of immune status in NSCLC immunotherapy, including MLR, NLR, NMLR, and PLR ^58,59^. These ratios reflect the balance between inflammatory and adaptive immune response ^60^. Within our cohort, NLR, NMLR, and PLR were elevated in NR patients, with PLR showing the strongest positive correlation with MUC1, MUC5B, MUC5AC, and ANPEP. This finding supports a link between these four proteins and systemic immune suppression. In addition, we developed a composite risk score integrating these four proteins with PLR. The score demonstrated robust discrimination of response to treatment, and was significantly associated with both PFS and OS. These results highlight the potential of an integrated EVs protein- and immune markers-based score to refine patient stratification and guide therapeutic decision-making in NSCLC.

However, this study has several limitations. First, the sample size is relatively small for a bioinformatics analysis of this nature. To mitigate this, we applied resampling strategies and machine learning approaches, including Random Forest modeling, which consistently highlighted the same proteins as robust predictors, reinforcing the validity of our findings. Moreover, these findings were experimentally validated by PRM, further supporting the robustness of the identified protein signatures. Second, the lack of an external cohort prevents formal assessment of scalability to new populations, highlighting the need for validation in larger and multicenter studies. In addition, the absence of standardized protocols for EVs isolation and clinical implementation currently limits the translational applicability of EVs-based biomarkers. It should be noted that, throughout this study, the term EVs refers to small extracellular vesicles (<200 nm) enriched for CD9/CD63, as their precise biogenetic origin cannot be unequivocally determined with current isolation methods ^61^. Finally, the cohort was predominantly male, and further studies including more women are required to ensure generalizability. Functional studies will also be essential to define the mechanistic role of these proteins in shaping the immunosuppressive tumor microenvironment and to explore potential synergistic effects interactions.

In conclusion, this study provides the first comprehensive analysis of the EV proteome in NSCLC patients treated with pembrolizumab. We identified MUC1, MUC5B, MUC5AC, and ANPEP as promising biomarkers of response to pembrolizumab, and propose a risk score integrating these proteins with PLR that significantly stratifies patient outcomes. These findings highlight the potential of an EVs protein–based panel, in combination with immune-derived markers, to advance precision medicine in NSCLC through the development of predictive and prognostic strategies.

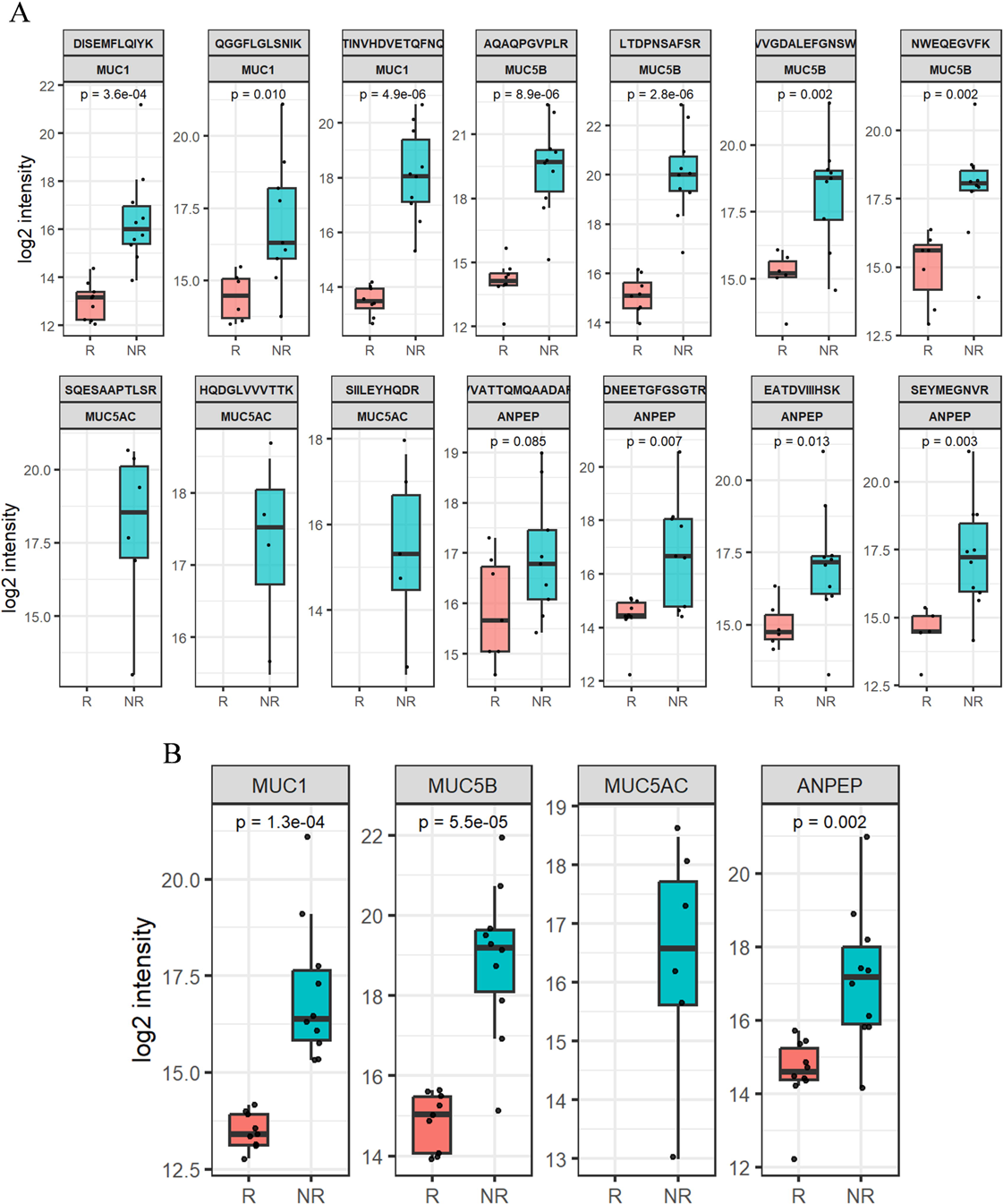

## Acknowledgments

We would like to acknowledge patients and their families for donating biological samples.

## Data availability statement

The mass spectrometry proteomics data have been deposited to the ProteomeXchange Consortium via the PRIDE ^62^ partner repository with the dataset identifier PXD071750.

