## Supplemental files for "Extracellular vesicles proteomics-based machine-learning model predicts immunotherapy response in NSCLC"

**Supplementary Table**

**Table S1. Peptide sequences and analytical parameters used in the PRM assay.**

| **Protein** | **Peptide** | **NR samples** | **R samples** | **Charge** | **m/z** | **Retention time (min)** |
| --- | --- | --- | --- | --- | --- | --- |
| MUC1 | DISEMFLQIYK | 9 | 10 | 2 | 693.85 | 73.257 |
| MUC1 | QGGFLGLSNIK | 9 | 6 | 2 | 567.31 | 62.800 |
| MUC1 | EGTINVHDVETQFNQYK | 10 | 8 | 3 | 674.657 | 55.960 |
| MUC5B | AQAQPGVPLR | 10 | 8 | 2 | 518.791 | 39.639 |
| MUC5B | LTDPNSAFSR | 10 | 8 | 2 | 554.275 | 39.912 |
| MUC5B | SVVGDALEFGNSWK | 9 | 6 | 2 | 754.870 | 68.604 |
| MUC5B | NWEQEGVFK | 10 | 7 | 2 | 568.770 | 50.925 |
| MUC5AC | HQDGLVVVTTK | 4 | 0 | 2 | 598.835 | 41.119 |
| MUC5AC | SQESAAPTLSR | 6 | 0 | 2 | 651.841 | 29.083 |
| MUC5AC | SIILEYHQDR | 5 | 0 | 2 | 637.330 | 45.231 |
| ANPEP | VVATTQMQAADAR | 10 | 7 | 2 | 681.345 | 35.266 |
| ANPEP | DNEETGFGSGTR | 10 | 9 | 2 | 699.318 | 26.339 |
| ANPEP | EATDVIIIHSK | 10 | 6 | 2 | 613.340 | 45.612 |
| ANPEP | SEYMEGNVR | 10 | 5 | 2 | 542.740 | 32.176 |

**Supplementary figures**

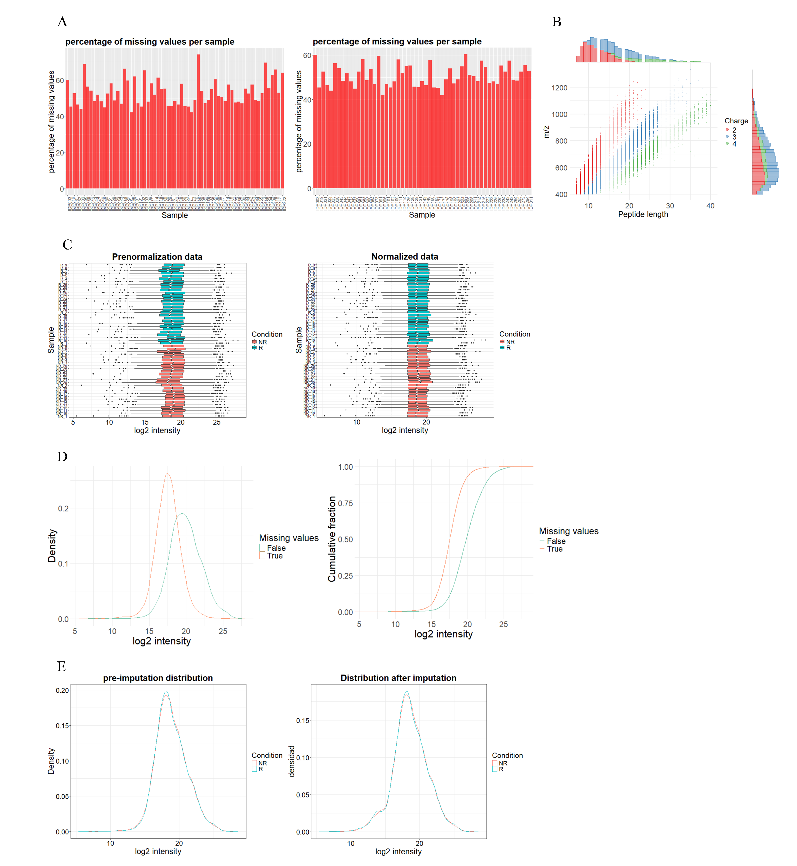
**Figure S1**

**Figure S2**

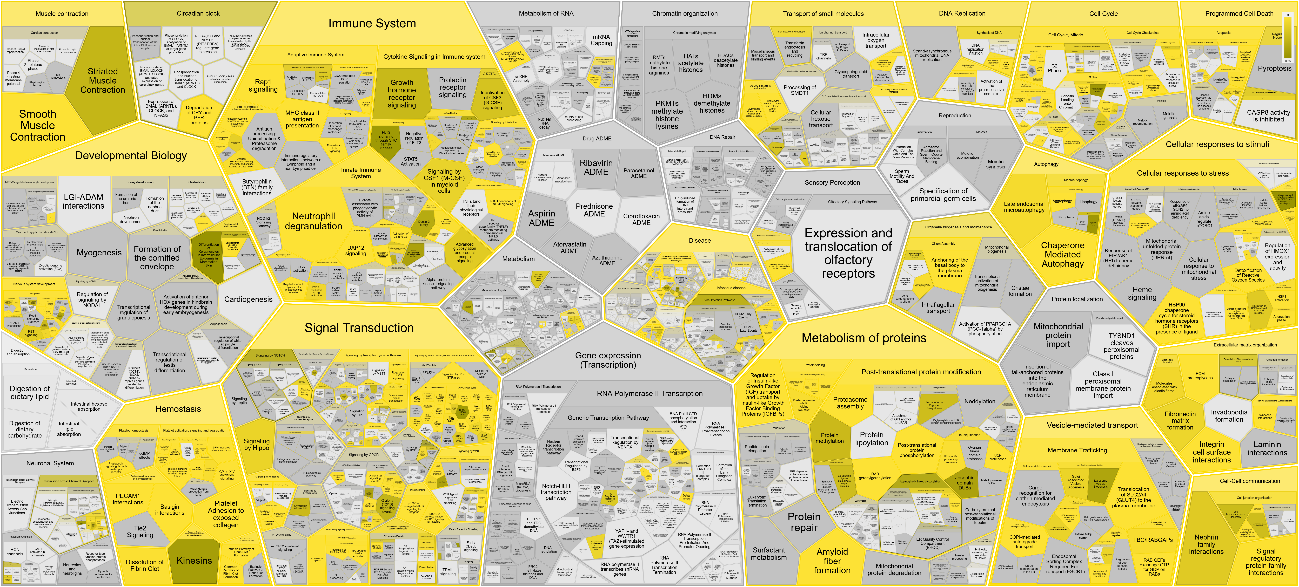

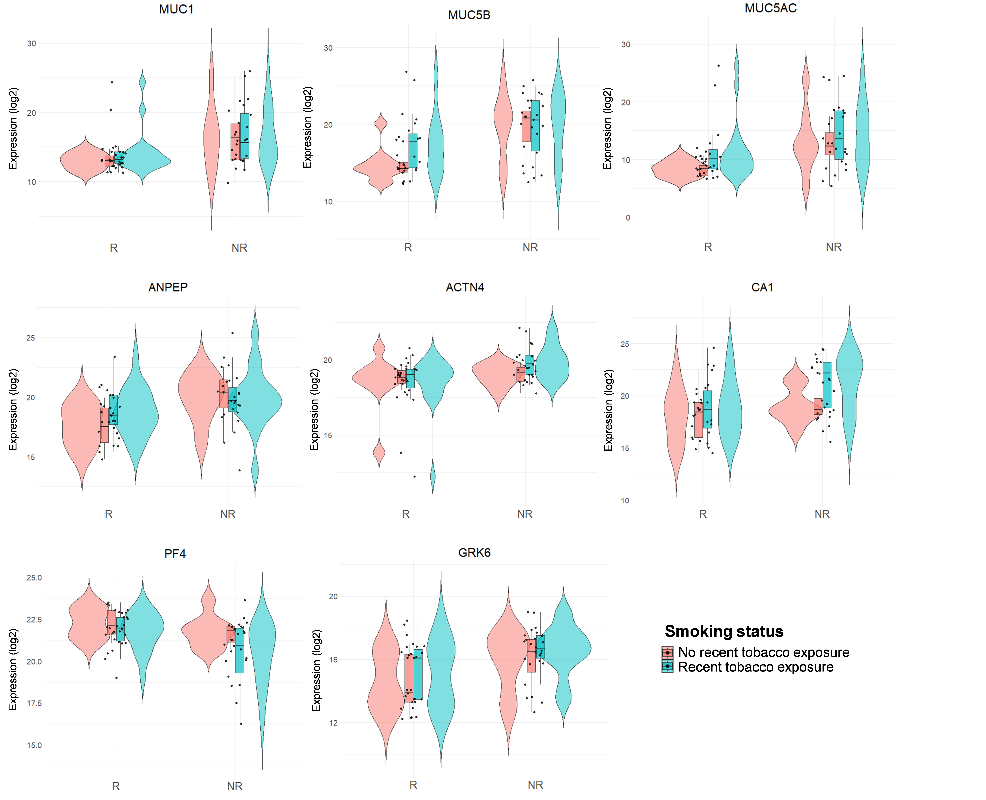
**Figure S3**

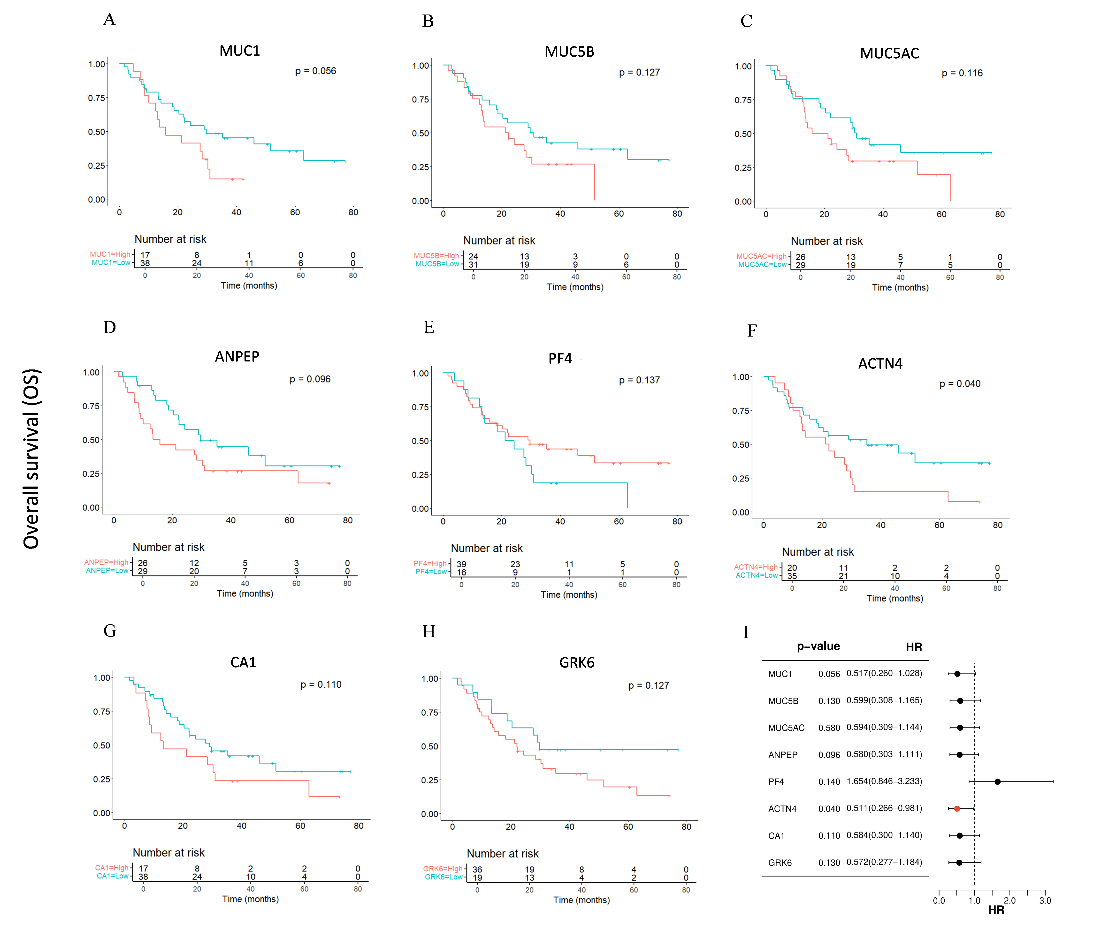
**Figure S4**

**Figure S5**

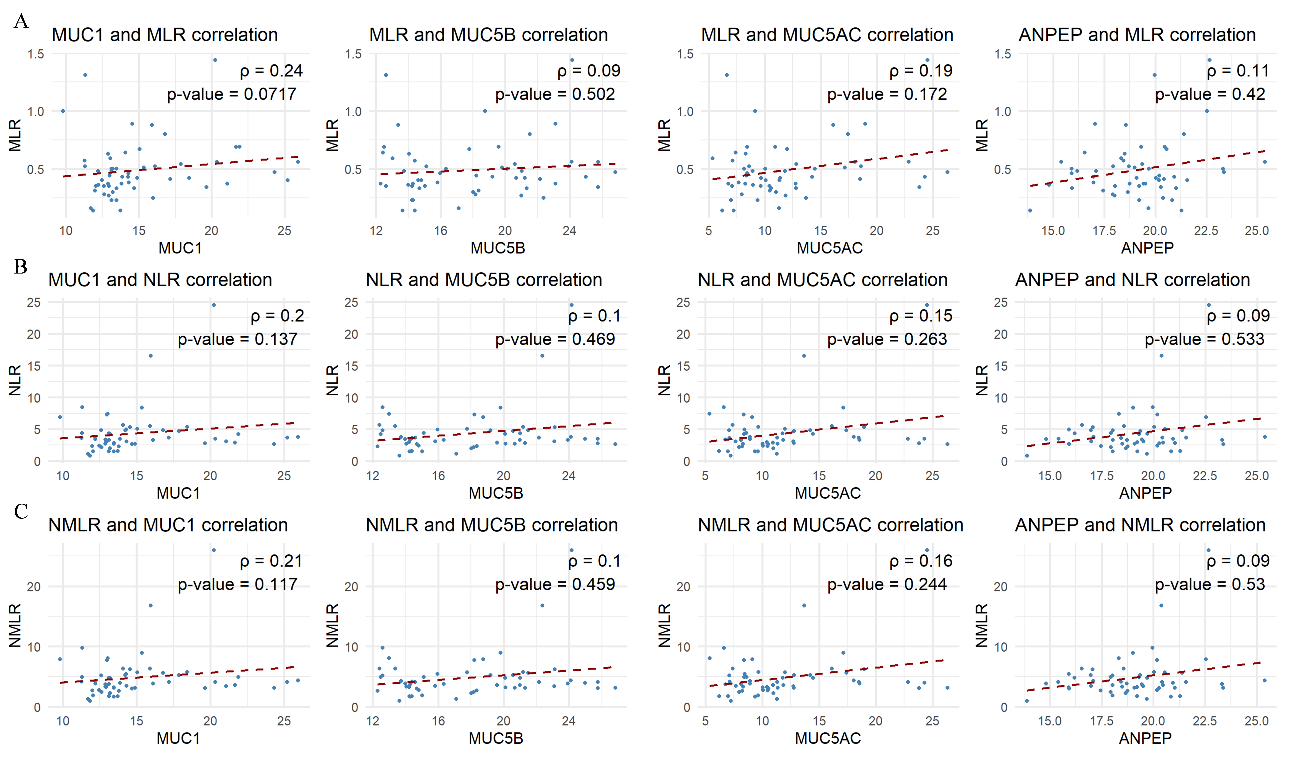

**Figure S6**

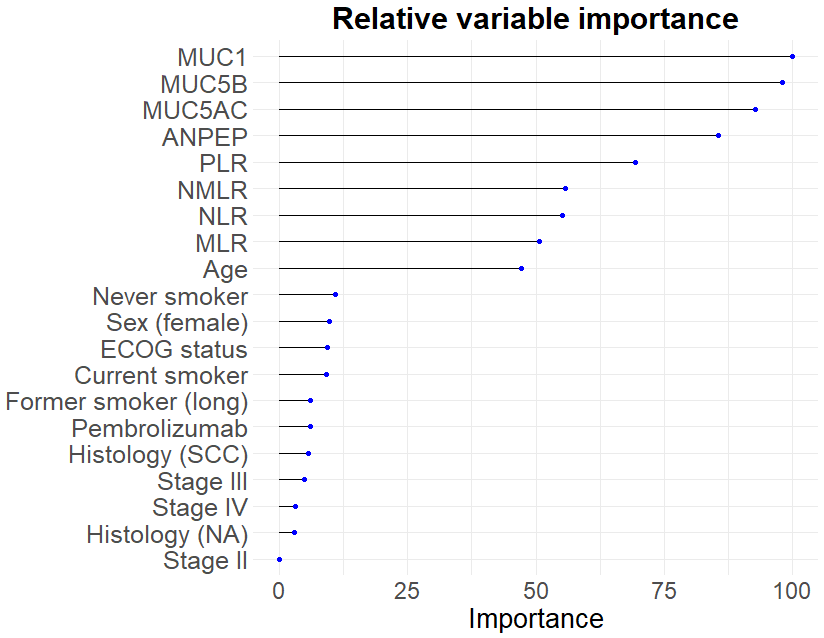

**Supplementary figure legends**

**Figure S1. Quality control of proteomic data processing. A)** Percentage of proteins with missing values per sample, before (left) and after (right) filtering samples with more than 60% missing values. **B)** Distribution of identified peptides by length and mass-to-charge ratio (m/z), color-coded according to charge state. Marginal histograms show peptide density by length (top) and by m/z (right). **C)** Boxplot of intensity distributions across samples before (left) and after (right) variance stabilization normalization. Color indicate the experimental group (NR in red; R in blue). **D)** Distribution of proteins intensities with missing values (red) vs. complete data (blue), shown as density (left) and cumulative fraction (right). Missing values concentrated in low-intensity proteins. **E)** Global intensity distributions by condition (NR in red; R in blue) before (left) and after (right) imputation. No major distortions were observed following the selected imputation strategy.

**Figure S2. Enriched biological pathways identified with Reactome.** Each cell represents a pathway organized hierarchically. Color intensity (yellow) indicates the degree of enrichment, while grey denotes pathways not reaching statistical significance.

**Figure S3. Protein expression stratified by treatment response and smoking status.** Violin-box plots show log2-transformed protein intensities for each candidate protein in R and NR, further stratified by smoking status. Each dot represents an individual sample and violins depicts the distribution density.

**Figure S4. Overall survival (OS) analysis of differentially expressed exosomal proteins. A-H)** Kaplan–Meier curves show OS stratified by high and low protein expression, with cut-off points determined using the Youden index. **A)** MUC1, **B)** MUC5B, **C)** MUC5AC, **D)** ANPEP, **E)** PF4, **F)** ACTN4, **G)** CA1, **H)** GRK6. P-values are indicated. **I)** Forest plot showing the hazard ratios (HR) and 95% confidence intervals derived from analyzed exosomal proteins. Red dots correspond to significant associations with HR < 1, blue dots to HR > 1, and black dots to proteins whose confidence interval includes HR = 1.

**Figure S5. Spearman correlation between the four top-ranked predictive proteins (MUC1, MUC5B, MUC5AC, and ANPEP) and A) MLR, B) NLR, C) and NMLR ratios.**

**Figure S6. Relevance of MUC1, MUC5B, MUC5AC, and ANPEP proteins in predicting response to immunotherapy.** Relative importance of the four proteins included in the final model compared with the clinical variables analyzed in the study, calculated using the Gini impurity metric. A Random Forest model with repeated k-fold cross-validation was applied. Only the top 20 variables are shown.
